# Menstrual symptoms and infertility odds among individuals with co-occurring endometriosis and polyendocrine metabolic ovarian syndrome

**DOI:** 10.64898/2026.10.13.26365449

**Authors:** Sophia Werdin, Zifan Wang, Eliana Huffman, Anne Marie Z Jukic, Allen J Wilcox, Sanaa Suharwardy, Jukka-Pekka Onnela, Brent A Coull, Michelle A Williams, Shruthi Mahalingaiah

**Author notes:** **Corresponding author:** Sophia Werdin, PhD, Department of Environmental Health, Harvard T.H. Chan School of Public Health, 665 Huntington Avenue, Boston, MA 02115, USA. **CRediT Authorship Contribution Statement:** Sophia Werdin: Conceptualization, Formal analysis, Investigation, Methodology, Visualization, Writing – original draft, Writing – review & editing; Zifan Wang: Writing – review & editing; Eliana Huffman: Writing – review & editing; Anne Marie Z Jukic: Writing – review & editing; Allen J Wilcox: Writing – review & editing; Sanaa Suharwardy: Writing – review & editing; Jukka-Pekka Onnela: Supervision, Writing – review & editing; Brent A Coull: Supervision, Writing – review & editing; Michelle A Williams: Supervision, Writing – review & editing; Shruthi Mahalingaiah: Conceptualization, Funding acquisition, Investigation, Methodology, Resources, Supervision, Writing – review & editing. **Attestation statements:** Data regarding any of the subjects in the study has not been previously published unless specified. Data will be made available to the editors of the journal for review or query upon request. **Data sharing statement:** Aggregated data that support the findings of this study may be available upon reasonable request to the corresponding author. Any request for data will be evaluated and responded to in a manner consistent with policies intended to protect participant confidentiality and language in the study protocol and informed consent form. **Capsule:** Co-occurring endometriosis and polyendocrine metabolic ovarian syndrome affected 1 in 50 participants and was associated with supra-additive infertility risk and poorer health, suggesting a distinct subpopulation needing tailored multidisciplinary care.

## Abstract

**Objective:** To study the prevalence of co-occurring endometriosis and polyendocrine metabolic ovarian syndrome (PMOS), estimate infertility odds, and characterize menstrual symptom profiles associated with this combined phenotype in a population cohort.

**Design:** Prospective digital cohort in the United States (US).

**Subjects:** 90,714 consented participants enrolled between November 2019 and June 2025 who completed relevant surveys; a subset of 25,152 participants with menstrual cycle logs linked to monthly surveys contributed to symptom analyses.

**Exposure:** Participants were classified by self-reported physician diagnosis into four groups: endometriosis alone, PMOS alone, co-occurring endometriosis and PMOS (endometriosis+PMOS), or neither condition (reference group).

**Main Outcome Measures:** (1) Odds of self-reported physician-diagnosed infertility (ever/never, cross-sectional), estimated using multivariable logistic regression adjusted for age, race/ethnicity, and subjective socioeconomic status; (2) number of adverse menstrual symptoms logged per cycle, assessed using negative binomial mixed-effects models adjusted for age, race/ethnicity, and hormonal contraceptive use.

**Results:** Among 90,714 participants, the mean age was 35.1 years (standard deviation: 11.6) and the majority identified as non-Hispanic White (72%). Prevalence estimates were 11% for PMOS alone, 6% for endometriosis alone, and 2% for endometriosis+PMOS. Individuals with endometriosis+PMOS more frequently reported unemployment, low socioeconomic status, active smoking, poorer subjective health, early menarche, current irregular cycles, and a broad range of comorbidities. Infertility odds were highest for endometriosis+PMOS (odds ratio: 12.49 [95% confidence interval: 11.13–13.99]), followed by PMOS alone (6.12 [5.69–6.59]) and endometriosis alone (4.31 [3.93–4.72]), with the combined effect exceeding additive expectations (relative excess risk due to interaction: 3.05 [1.65–4.46]). Individuals with endometriosis+PMOS logged 11% more symptoms per cycle than those with neither condition (incidence rate ratio: 1.11 [1.04–1.19]) and had the highest odds of logging pelvic pain, fatigue, and sleep changes, among other symptoms.

**Conclusion:** In this large US digital cohort, co-occurring endometriosis and PMOS affected 1 in 50 participants and was associated with markedly elevated infertility odds exceeding additive expectations, and a heightened menstrual symptom burden. Several risk factors and health impacts characteristic of each condition appeared amplified in individuals with both conditions, suggesting a subgroup with compounded clinical complexity and greater care needs.

## Introduction

Endometriosis and polyendocrine metabolic ovarian syndrome (PMOS; the updated name for polycystic ovary syndrome proposed in 2026 (1)) are among the most prevalent chronic multisystem conditions affecting reproductive-age individuals. While each condition is underpinned by a distinct and complex pathophysiology, both are leading causes of female infertility (2) and share risk factors and long-term disease outcomes (3). PMOS affects an estimated 8–13% of menstruating individuals globally, with prevalence varying by diagnostic criteria (4). Driven by dysregulation of neuroendocrine, insulin, androgen, and ovarian hormone pathways, PMOS may manifest metabolic, reproductive, cardiovascular, psychological, and dermatologic features (5–7). Common symptoms include infrequent or prolonged menstrual cycles, hirsutism, and acne (1). Endometriosis affects approximately 10% of reproductive-age women, is characterized by the growth of endometrial-like tissue outside the uterus, and commonly presents with chronic pelvic pain, dysmenorrhea, and dyspareunia (8). Both conditions are substantially underdiagnosed, with diagnostic delays often spanning years attributable to factors including normalization of symptoms, the complexity of diagnostic criteria, and limited awareness among patients and clinicians alike (9–11).

PMOS and endometriosis are often considered unrelated or diametric disorders (12–15). While both conditions are thought to arise from alterations in prenatal testosterone exposure and dysregulation of the hypothalamic-pituitary-ovarian axis, they occupy opposite ends of that spectrum (13,15–18). In PMOS, elevated gonadotropin-releasing hormone pulse frequency is thought to drive a luteinizing hormone-dominant pattern, promoting follicular arrest, anovulation, and an androgen-dominant hormonal environment (19–21). Conversely, in endometriosis, reduced gonadotropin-releasing hormone pulse frequency is thought to cause a follicle-stimulating hormone-dominant pattern, favoring follicular growth, ovulation, and an estrogen-dominant hormonal environment (13,22–24). These opposing disturbances in steroid balance shape fundamentally different patterns of ovarian function and distinct hormonal milieus in each condition (12), and their divergent pathophysiology suggests that co-occurrence within the same individual would be unlikely.

However, a growing body of evidence has documented the biologically unique phenotype of co-occurring endometriosis and PMOS and has begun to characterize the clinical profile of affected individuals (3,25). In a US operative cohort, Schliep et al. (3) found that patients with co-occurring endometriosis and PMOS had the highest rates of subfertility and chronic pelvic pain compared to those with one condition alone or neither condition. An analysis of a US inpatient sample database further demonstrated that individuals with co-occurring endometriosis and PMOS had higher rates of pregnancy complications than those with PMOS alone (14). Despite these emerging findings, evidence on the combined phenotype remains largely derived from small clinical samples focused on individuals with infertility, with limited research examining its broader health effects. While menstrual symptoms and infertility risk have been well-characterized in each condition individually, estimates of their combined effects in population-based samples have yet to be established. Generating such insights could help inform clinical recognition of this unique phenotype, reduce diagnostic delay, and guide more tailored management strategies for individuals presenting with features of both conditions. Accordingly, this study aimed to quantify the prevalence of co-occurrence, estimate infertility odds, and characterize menstrual symptom profiles among individuals with co-occurring endometriosis and PMOS in a large, digital US cohort.

## Materials and Methods

### Study design and population

We analyzed self-reported health data from the Apple Women’s Health Study (AWHS), a large, prospective, US-based digital cohort enrolling participants since November 2019. Participants must reside in the US, be at least 18 years old (19 in Alabama and Nebraska, 21 in Puerto Rico), have menstruated at least once, be the sole user of an iPhone/iCloud account, be able to communicate in English, and provide informed consent (26). All participants actively enrolled in the study and explicitly opted in to share their data for research purposes. Detailed study procedures have been described previously (26). The AWHS was approved by the Institutional Review Board at Advarra (CIRB #PRO00037562) and registered on ClinicalTrials.gov (NCT04196595).

At enrollment, participants completed surveys on demographics, medical and reproductive history, and lifestyle behaviors, with monthly follow-up surveys collecting contextual data on the previous menstrual cycle, including hormonal contraceptive use (26). Participants could also link mobile menstrual cycle logs and clinical health records (CHR) to the study. For this analysis, menstrual cycle logging data were restricted to the Apple Health app, where standardized symptom categories were consistently available (27). We included participants enrolled through June 9, 2025, who completed baseline surveys on demographics and medical history. For symptom analyses, we further restricted the sample to participants who did not report a history of hysterectomy or bilateral oophorectomy, logged at least one menstrual symptom through March 1, 2025, and completed the corresponding monthly survey (Supplemental figure 1).

### Sociodemographic characteristics, medical history, and hormonal contraceptive use

Sociodemographic characteristics and medical history were self-reported at enrollment, including age, race/ethnicity, subjective socioeconomic status (SES), body mass index (BMI; calculated from self-reported height and weight), and physician-diagnosed gynecologic conditions, including infertility (“Have you ever been diagnosed with any of the following by a doctor or other care provider?”). Subjective SES was assessed using the MacArthur Scale of Subjective Social Status, a validated single-item social ladder measure in which participants ranked themselves relative to others (28). Parity was defined as the number of self-reported pregnancies lasting ≥24 weeks of gestation. Participants were classified into four mutually exclusive groups: endometriosis alone, PMOS alone, co-occurring endometriosis and PMOS, or neither condition. As data collection preceded the PMOS nomenclature update (1), participants self-reported a physician diagnosis of polycystic ovary syndrome. Throughout this paper, polycystic ovary syndrome diagnoses have been mapped to the updated PMOS label. To validate self-reported diagnoses, we evaluated a subset of participants who shared their CHR data and assessed condition co-occurrence in this clinically confirmed subset. In symptom analyses, age at symptom logging was used rather than baseline age. Both age at symptom logging and hormonal contraceptive use were defined at the cycle level to capture within-person variation.

### Menstrual cycles and symptoms

Symptom analyses included bleeding days and symptoms logged in the Apple Health app. Each of 23 prespecified adverse symptoms could be logged once per day, yielding binary day-level indicators. Only post-enrollment entries linked to logged bleeding dates and corresponding monthly survey responses were included. A menstrual cycle was defined as one or more consecutive bleeding days followed by at least two non-bleeding days, with the first bleeding day marking cycle onset (29). Cycles shorter than 10 or longer than 90 days were excluded as unlikely to represent natural cycles (30).

### Statistical analyses

Baseline characteristics were summarized descriptively. Infertility odds across condition groups were estimated using multivariable logistic regression adjusted for age, race/ethnicity, and subjective SES. To account for nonlinear associations between age and infertility, age was modeled using natural cubic splines with three interior knots (at the 25^th^, 50^th^, and 75^th^ percentiles), with the number of knots selected based on Akaike Information Criterion comparison. As a secondary analysis, the model was reparameterized using two binary indicators for endometriosis and PMOS status and their interaction term to decompose the main effect of each condition and assess additive interaction via the relative excess risk due to interaction (RERI = OR_Endometriosis+PMOS_ – OR_Endometriosis alone_ – OR_PMOS alone_ + 1). A further secondary analysis stratified infertility by baseline parity, distinguishing primary (parity = 0) from secondary infertility (parity ≥1). Since infertility diagnosis and parity were both assessed at baseline, temporal ordering could not be established and related findings should be considered exploratory.

Menstrual-symptom prevalence was defined as the proportion of participants logging each symptom at least once across all included cycles. For each menstrual symptom, univariable mixed-effects logistic regression with a subject-specific random intercept estimated odds of logging that symptom in a cycle by condition group. Overall menstrual symptom burden per cycle was examined using negative binomial mixed-effects regression with a subject-specific random intercept and a log link to account for overdispersed count data, adjusting for condition group, age at symptom logging, race/ethnicity, and hormonal contraceptive use. Individuals with neither condition served as the reference in all analyses. Given minimal missing data across survey variables (≤2% per variable), complete case analyses were performed. Data analysis was performed in R version 4.1.2.

## Results

### Baseline sociodemographic characteristics

The primary analytic sample comprised 90,714 participants with a mean age of 35.1 years (SD = 11.6; Table 1). Most identified as non-Hispanic White (72%), were highly educated (28% college degree, 22% graduate-level education), and were employed (72%). Endometriosis alone was reported by 6% (n = 5,635), PMOS alone by 11% (n = 9,695), and endometriosis+PMOS by 2% (n = 1,872) (Supplemental figure 2). Infertility was reported by 5% (n = 4,676). Endometriosis alone had the highest mean age (40.5 years, SD = 12.2), while PMOS alone had the lowest (33.8 years, SD = 9.2). Endometriosis alone and endometriosis+PMOS had the highest proportions of non-Hispanic White individuals (78% and 76%, respectively) and highest rates of unemployment or inability to work (15% and 18%, respectively), while subjective SES was lowest in the endometriosis+PMOS group. PMOS alone had the highest prevalence of obesity (60%), followed by endometriosis+PMOS (54%) and endometriosis alone (36%).

**Table 1.**
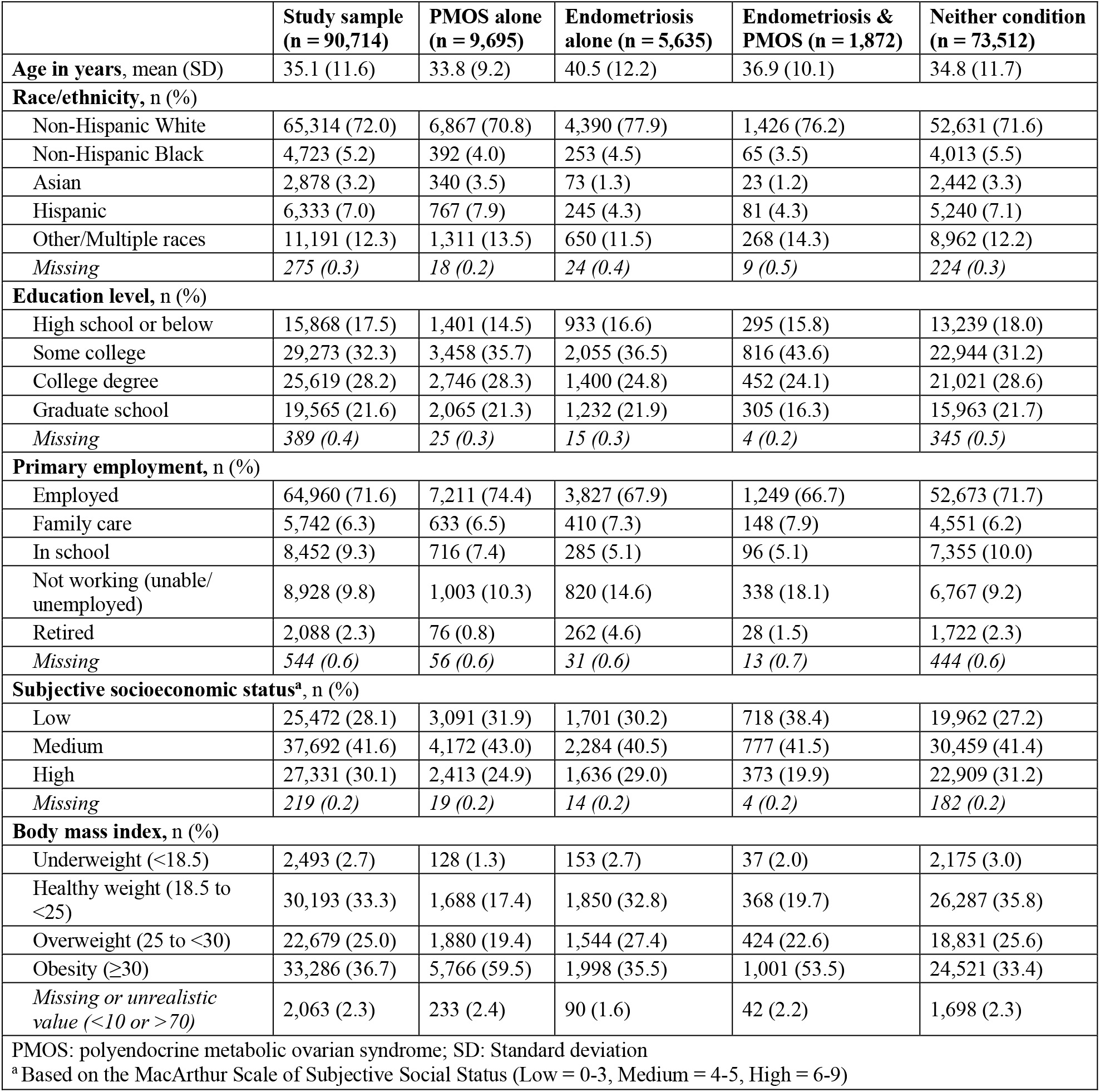
Demographic and anthropometric characteristics at baseline by condition group.

|  | **Study sample**<br>**(n = 90,714)** | **PMOS alone**<br>**(n = 9,695)** | **Endometriosis**<br>**alone (n = 5,635)** | **Endometriosis &**<br>**PMOS (n = 1,872)** | **Neither condition**<br>**(n = 73,512)** |
| --- | --- | --- | --- | --- | --- |
| **Age in years, mean (SD)** | 35.1 (11.6) | 33.8 (9.2) | 40.5 (12.2) | 36.9 (10.1) | 34.8 (11.7) |
| **Race/ethnicity, n (%)** |  |  |  |  |  |
| Non-Hispanic White | 65,314 (72.0) | 6,867 (70.8) | 4,390 (77.9) | 1,426 (76.2) | 52,631 (71.6) |
| Non-Hispanic Black | 4,723 (5.2) | 392 (4.0) | 253 (4.5) | 65 (3.5) | 4,013 (5.5) |
| Asian | 2,878 (3.2) | 340 (3.5) | 73 (1.3) | 23 (1.2) | 2,442 (3.3) |
| Hispanic | 6,333 (7.0) | 767 (7.9) | 245 (4.3) | 81 (4.3) | 5,240 (7.1) |
| Other/Multiple races | 11,191 (12.3) | 1,311 (13.5) | 650 (11.5) | 268 (14.3) | 8,962 (12.2) |
| *Missing* | 275 (0.3) | 18 (0.2) | 24 (0.4) | 9 (0.5) | 224 (0.3) |
| **Education level, n (%)** |  |  |  |  |  |
| High school or below | 15,868 (17.5) | 1,401 (14.5) | 933 (16.6) | 295 (15.8) | 13,239 (18.0) |
| Some college | 29,273 (32.3) | 3,458 (35.7) | 2,055 (36.5) | 816 (43.6) | 22,944 (31.2) |
| College degree | 25,619 (28.2) | 2,746 (28.3) | 1,400 (24.8) | 452 (24.1) | 21,021 (28.6) |
| Graduate school | 19,565 (21.6) | 2,065 (21.3) | 1,232 (21.9) | 305 (16.3) | 15,963 (21.7) |
| *Missing* | 389 (0.4) | 25 (0.3) | 15 (0.3) | 4 (0.2) | 345 (0.5) |
| **Primary employment, n (%)** |  |  |  |  |  |
| Employed | 64,960 (71.6) | 7,211 (74.4) | 3,827 (67.9) | 1,249 (66.7) | 52,673 (71.7) |
| Family care | 5,742 (6.3) | 633 (6.5) | 410 (7.3) | 148 (7.9) | 4,551 (6.2) |
| In school | 8,452 (9.3) | 716 (7.4) | 285 (5.1) | 96 (5.1) | 7,355 (10.0) |
| Not working (unable/<br>unemployed) | 8,928 (9.8) | 1,003 (10.3) | 820 (14.6) | 338 (18.1) | 6,767 (9.2) |
| Retired | 2,088 (2.3) | 76 (0.8) | 262 (4.6) | 28 (1.5) | 1,722 (2.3) |
| *Missing* | 544 (0.6) | 56 (0.6) | 31 (0.6) | 13 (0.7) | 444 (0.6) |
| **Subjective socioeconomic status<sup>a</sup>, n (%)** |  |  |  |  |  |
| Low | 25,472 (28.1) | 3,091 (31.9) | 1,701 (30.2) | 718 (38.4) | 19,962 (27.2) |
| Medium | 37,692 (41.6) | 4,172 (43.0) | 2,284 (40.5) | 777 (41.5) | 30,459 (41.4) |
| High | 27,331 (30.1) | 2,413 (24.9) | 1,636 (29.0) | 373 (19.9) | 22,909 (31.2) |
| *Missing* | 219 (0.2) | 19 (0.2) | 14 (0.2) | 4 (0.2) | 182 (0.2) |
| **Body mass index, n (%)** |  |  |  |  |  |
| Underweight (<18.5) | 2,493 (2.7) | 128 (1.3) | 153 (2.7) | 37 (2.0) | 2,175 (3.0) |
| Healthy weight (18.5 to<br><25) | 30,193 (33.3) | 1,688 (17.4) | 1,850 (32.8) | 368 (19.7) | 26,287 (35.8) |
| Overweight (25 to <30) | 22,679 (25.0) | 1,880 (19.4) | 1,544 (27.4) | 424 (22.6) | 18,831 (25.6) |
| Obesity (≥30) | 33,286 (36.7) | 5,766 (59.5) | 1,998 (35.5) | 1,001 (53.5) | 24,521 (33.4) |
| *Missing or unrealistic*<br>*value (<10 or >70)* | 2,063 (2.3) | 233 (2.4) | 90 (1.6) | 42 (2.2) | 1,698 (2.3) |
| PMOS: polyendocrine metabolic ovarian syndrome; SD: Standard deviation |  |  |  |  |  |
| <sup>a</sup> Based on the MacArthur Scale of Subjective Social Status (Low = 0-3, Medium = 4-5, High = 6-9) |  |  |  |  |  |

Of the 90,714 participants in the primary analytic sample, 17,165 individuals (19%) contributed CHR data. In this subsample, self-reported prevalences were consistently higher than clinically confirmed diagnoses (endometriosis alone: 7% vs. 2%, PMOS alone: 12% vs. 6%, endometriosis+PMOS: 3% vs. 0.3%, infertility: 6% vs. 1%).

### Reproductive history, medical history, and lifestyle behaviors

Endometriosis+PMOS had the highest prevalence of nearly all adverse reproductive characteristics and comorbidities examined (Figure 1). Early menarche, irregular cycles, and hormone use for menstrual-cycle management and fertility were most common with endometriosis+PMOS and PMOS alone. The combined group and endometriosis alone had the highest prevalence of other gynecologic conditions and surgeries. This pattern extended to broader medical history, with the endometriosis+PMOS group having particularly elevated rates of chronic fatigue syndrome (19% vs. 4%), pelvic inflammatory disease (12% vs. 2%), and fibromyalgia (19% vs. 4%) relative to individuals with neither condition. Participants with endometriosis+PMOS also reported the poorest subjective health, the highest frequency of feeling overly sleepy during the day, and the highest rates of active smoking (Supplemental table 1).

**Figure 1.**
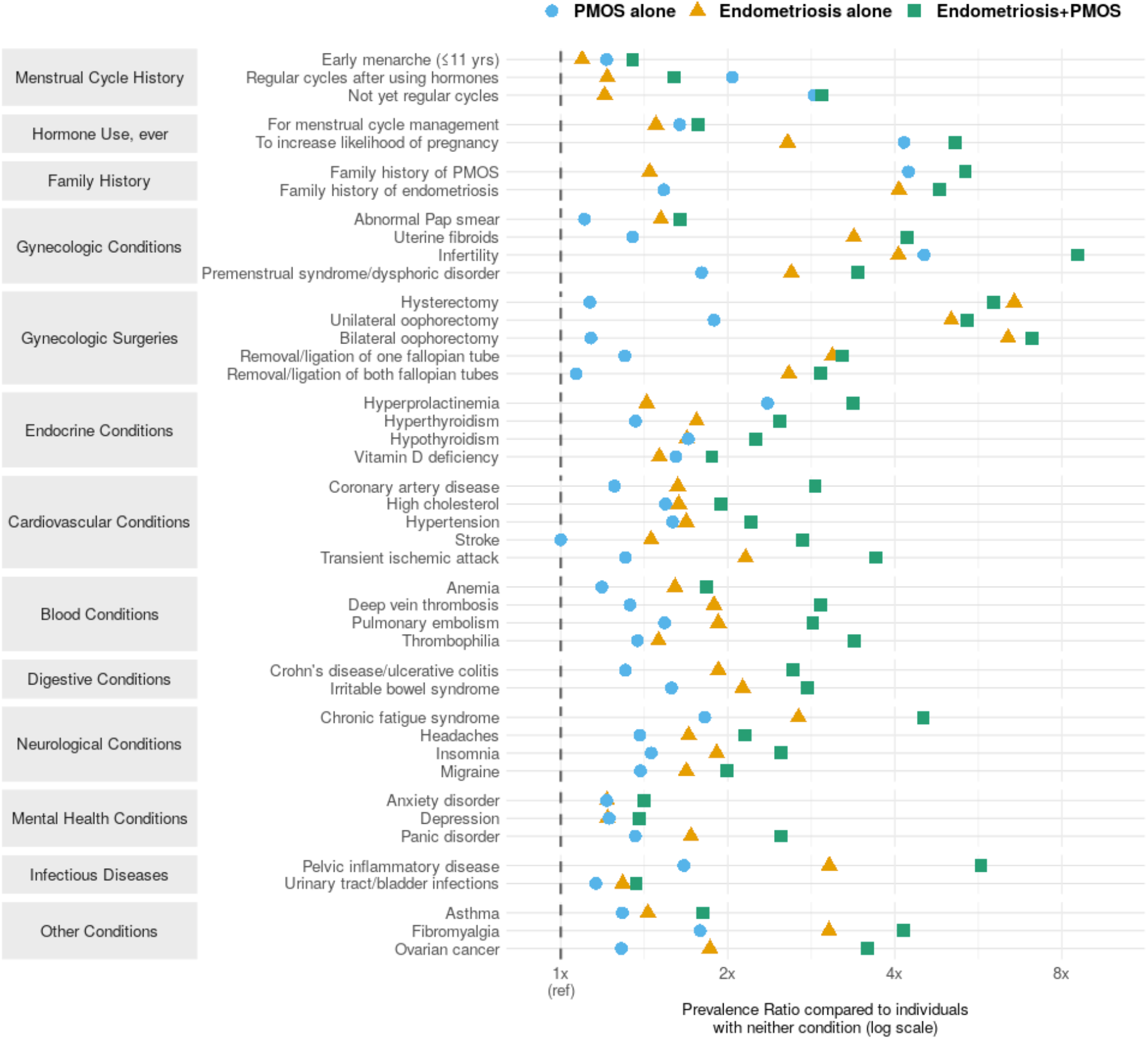
Prevalence ratios for reproductive history, gynecologic conditions and surgeries, and other health conditions by condition group, relative to individuals with neither condition (reference group indicated by dashed line at 1x). The x-axis is displayed on a log scale. PMOS = polyendocrine metabolic ovarian syndrome; Endometriosis+PMOS = co-occurring endometriosis and PMOS.

### Association between condition groups and infertility

After adjusting for age, race/ethnicity, and subjective SES, all three condition groups were associated with significantly elevated infertility odds compared to those with neither condition (Table 2). Infertility odds were highest for endometriosis+PMOS (OR = 12.49, 95% CI: 11.13–13.99), followed by PMOS alone (OR = 6.12, 95% CI: 5.69–6.59) and endometriosis alone (OR = 4.31, 95% CI: 3.93–4.72). The combined effect exceeded additive expectations (RERI = 3.05, 95% CI: 1.65–4.46), indicating supra-additive interaction. Among covariates, higher subjective SES and non-Hispanic White race/ethnicity were associated with increased infertility odds. In the parity-stratified analysis, 2,032 infertile participants (44%) were classified as having primary and 2,484 (53%) as secondary infertility; 160 participants (3%) could not be classified due to missing parity data. The pattern of elevated infertility odds was consistent across subtypes, with endometriosis+PMOS showing the highest odds for both primary (OR = 14.19, 95% CI: 11.87–16.92) and secondary infertility (OR = 11.34, 95% CI: 9.69–13.24).

**Table 2.** Multivariable logistic regression analysis of infertility odds by gynecologic condition group, including additive interaction decomposition for co-occurring endometriosis and polyendocrine metabolic ovarian syndrome.

|  | OR | 95% CI | P-value |
| --- | --- | --- | --- |
| **Primary analysis (four-category exposure)** |  |  |  |
| **Condition group** (reference: Neither condition) |  |  |  |
| Polyendocrine metabolic ovarian syndrome alone | 6.12 | 5.69; 6.59 | <0.001 |
| Endometriosis alone | 4.31 | 3.93; 4.72 | <0.001 |
| Co-occurring endometriosis and polyendocrine metabolic ovarian syndrome | 12.49 | 11.13; 13.99 | <0.001 |
| **Baseline age in years** (modeled as splines, 3 interior knots) | - | - | - |
| **Race/ethnicity** (reference: Non-Hispanic White) |  |  |  |
| Non-Hispanic Black | 0.82 | 0.70; 0.95 | 0.009 |
| Asian | 0.58 | 0.46; 0.72 | <0.001 |
| Hispanic | 0.65 | 0.56; 0.75 | <0.001 |
| Other/multiple races/ethnicities | 0.91 | 0.82; 1.00 | 0.056 |
| **Subjective socioeconomic status** (reference: Low) |  |  |  |
| Medium | 1.16 | 1.07; 1.26 | <0.001 |
| High | 1.37 | 1.26; 1.49 | <0.001 |
| **Secondary analysis (interaction decomposition)** |  |  |  |
| Polyendocrine metabolic ovarian syndrome (main effect) | 6.12 | 5.69; 6.59 |  |
| Endometriosis (main effect) | 4.31 | 3.93; 4.72 |  |
| Endometriosis × Polyendocrine metabolic ovarian syndrome (interaction term) | 0.47 | 0.41; 0.55 |  |
| Endometriosis + Polyendocrine metabolic ovarian syndrome (joint effect) | 12.49 | 11.13; 13.99 |  |
| **Relative Excess Risk due to Interaction** | **3.05** | **1.65; 4.46** |  |
| Complete case analysis of 90,224 participants; those with missing data on any model covariate were excluded. Covariate estimates in the secondary analysis were identical to those in the primary analysis. OR: odds ratio, CI: confidence interval |  |  |  |

### Menstrual symptoms

The analytic sample for menstrual symptoms comprised 25,152 participants (PMOS alone: 2,775; endometriosis alone: 1,103; endometriosis+PMOS: 375; neither condition: 20,899), contributing 171,549 menstrual cycles and 648,255 symptom entries. The most commonly logged symptom was abdominal cramps, reported by 65% of participants at least once, followed by fatigue (42%), bloating (39%), and acne (39%). Logging of pelvic pain showed the largest effect sizes across condition groups, with the highest odds in the endometriosis+PMOS group (OR = 3.07, 95% CI: 2.32–4.05), followed by endometriosis alone (OR = 2.50, 95% CI: 2.12–2.95) and PMOS alone (OR = 1.38, 95% CI: 1.22–1.56). Additional symptoms with significantly higher odds of logging in the combined group compared to individuals with neither condition included chills, dry skin, hair loss, bladder incontinence, nausea, fatigue, and sleep changes (Supplemental table 2). After adjusting for age, race/ethnicity, and hormonal contraceptive use, all condition groups logged significantly more symptoms per cycle than participants with neither condition (Table 3). The increased symptom burden was comparable for endometriosis alone (IRR = 1.14, 95% CI: 1.09–1.18) and endometriosis+PMOS (IRR = 1.11, 95% CI: 1.04–1.19), and modestly smaller for PMOS alone (IRR = 1.06, 95% CI: 1.03–1.08).

**Table 3.** Negative binomial mixed-effects regression analysis of menstrual symptom burden per cycle by gynecologic condition group.

|  | IRR | 95% CI | P-value |
| --- | --- | --- | --- |
| **Condition group** (reference: Neither condition) |  |  |  |
| Polyendocrine metabolic ovarian syndrome alone | 1.06 | 1.03; 1.08 | <0.001 |
| Endometriosis alone | 1.14 | 1.09; 1.18 | <0.001 |
| Co-occurring endometriosis and polyendocrine metabolic ovarian syndrome | 1.11 | 1.04; 1.19 | 0.003 |
| **Age in years** | 0.99 | 0.99; 1.00 | <0.001 |
| **Race/ethnicity** (reference: Non-Hispanic White) |  |  |  |
| Non-Hispanic Black | 0.97 | 0.94; 1.01 | 0.171 |
| Asian | 0.95 | 0.91; 1.00 | 0.039 |
| Hispanic | 0.94 | 0.91; 0.97 | <0.001 |
| Other/multiple races/ethnicities | 1.02 | 0.99; 1.05 | 0.133 |
| **Hormone use in previous month** (reference: No) |  |  |  |
| Yes | 0.99 | 0.98; 1.01 | 0.372 |
| Complete case analysis; the model was fitted on 175,822 cycle-level observations from 25,085 participants. Those with missing data on any model covariate were excluded. IRR: incidence rate ratio, CI: confidence interval |  |  |  |

## Discussion

In this large US digital cohort of over 90,000 reproductive-age individuals, co-occurring endometriosis and PMOS affected 2% of participants and was associated with markedly elevated infertility odds, a distinct menstrual symptom pattern, and a broadly poorer health profile compared to those with either condition alone or neither condition. Infertility odds exceeded the sum of individual condition effects, indicating supra-additive interaction. Participants with endometriosis+PMOS had the highest prevalence of all examined health conditions, including chronic fatigue syndrome, fibromyalgia, and pelvic inflammatory disease, together with the poorest subjective health. Prospectively logged menstrual symptoms showed an elevated symptom burden in the combined group and the highest odds of logging pelvic pain, fatigue, and sleep changes, among other symptoms. Together, these findings indicate that individuals with comorbid endometriosis and PMOS may constitute a clinically distinct subpopulation facing a disproportionate health burden, underscoring the need for tailored diagnostic approaches and multidisciplinary management strategies.

Despite a considerably smaller population cohort of 127 women investigated by Schliep et al. (3), and slightly differing prevalence estimates for endometriosis alone (10% vs. 6%) and PMOS alone (8% vs. 11%), the prevalence estimate for co-occurring endometriosis+PMOS matched our finding of 2%. Other studies examining the prevalence of endometriosis among individuals with PMOS have reported wide-ranging estimates, including 1% among patients with PMOS who delivered (14), 8% among women with clomiphene citrate-resistant PMOS undergoing laparoscopic ovarian drilling (31), and 74% among women with PMOS and infertility or pelvic pain who underwent laparoscopy (25), reflecting substantial heterogeneity across study populations. In our population-based sample, 16% of individuals with self-reported PMOS also reported endometriosis - an estimate that, given the larger sample size, may provide a useful reference point within this wide range of clinical estimates.

Consistent with previous work (3,12,14,25), individuals with endometriosis+PMOS were more likely to identify as non-Hispanic White, had a lower mean BMI than those with PMOS alone but a higher BMI than those with endometriosis alone, and more frequently reported early menarche. The observed sociodemographic differences in disease and infertility prevalence by race/ethnicity and subjective SES may partly reflect differential healthcare access rather than true differences in disease burden, with unmeasured social determinants of health potentially contributing to underdiagnosis in certain groups (32,33). The intermediate BMI in the combined group may reflect competing metabolic influences, where insulin resistance, hyperandrogenism, and obesity characteristic of PMOS (34) may be partially offset by the lower BMI and altered metabolic signaling associated with endometriosis (35). Early menarche can be linked to greater lifetime exposure to ovarian hormones and has been associated with increased risk of endometriosis (36,37), with some studies also suggesting a link to PMOS (38,39). The higher prevalence of early menarche with endometriosis+PMOS may therefore reflect a shared hormonal vulnerability or cumulative risk effect, potentially accelerating the pathological processes underlying both conditions.

Our study extends previous work by demonstrating a broadly poorer health profile across reproductive and medical history domains, suggesting that co-occurrence may constitute a distinct and more severe phenotype with its own clinical trajectory. PMOS is a multisystem condition with well-established metabolic, reproductive, and psychological manifestations, including hypertension, cardiovascular disease, sleep apnea, depression, and anxiety (5,40). Consistent with this evidence, both PMOS groups showed elevated prevalence of these features compared to individuals with endometriosis alone or neither condition. Notably, the combined group showed an even greater burden of nearly all PMOS-associated multisystem features than the PMOS alone group, except for obesity and irregular menstrual cycle history. Rates of gynecologic surgeries, including hysterectomy and oophorectomy, were closer to those with endometriosis alone, suggesting that the surgical burden in co-occurring endometriosis+PMOS is driven predominantly by endometriosis. The higher burden of comorbidities may contribute to, and likely interacts with, the highest rates of unemployment, lowest subjective socioeconomic status, and poorest self-reported health observed in the combined group. Given well-documented diagnostic delays for endometriosis and PMOS individually (10,11), clinician awareness of the combined phenotype is particularly important to facilitate timely recognition and more intensive monitoring of patients presenting with features of both conditions. Our findings underscore the need for multidisciplinary care models that reflect the systemic health burden affected individuals face, with routine screening for common comorbidities, including fatigue-related disorders and pelvic inflammatory disease, particularly warranted.

In an operative cohort of 473 women, Schliep et al. (3) found that a history of subfertility was associated with a tenfold higher adjusted probability of having co-occurring endometriosis and PMOS compared with neither condition, which is broadly consistent with the eightfold higher unadjusted prevalence of infertility observed in the combined group in our sample. After adjusting for relevant covariates, we found 12-fold higher odds of infertility in the co-occurring group compared to individuals with neither condition, markedly exceeding the odds observed for either condition alone and surpassing additive expectations. These findings have direct clinical implications, including that proactive fertility counseling should be integrated into routine care for patients with both endometriosis and PMOS. This could include early discussions regarding fertility preservation strategies (e.g., oocyte cryopreservation), particularly for younger individuals who have not yet completed their reproductive plans but face a disproportionately high risk of infertility. Furthermore, individuals with co-occurring endometriosis and PMOS may be at risk of being evaluated and managed within a single diagnostic framework - either endometriosis or PMOS - rather than both. As each condition carries distinct diagnostic and therapeutic implications (41,42), recognizing co-occurrence may directly inform treatment selection and potentially improve assisted reproductive technology outcomes for those seeking fertility care, though dedicated research in this area is warranted.

In the menstrual symptom subsample of 25,152 participants, all condition groups were associated with a higher symptom burden compared to those with neither condition. Individuals with endometriosis+PMOS showed the highest odds of logging pelvic pain, chills, dry skin, hair loss, bladder incontinence, nausea, fatigue, and sleep changes, suggesting a distinct and broad symptom profile. The markedly elevated odds of logging pelvic pain with endometriosis+PMOS compared to endometriosis alone may reflect additive nociceptive or inflammatory mechanisms arising from the co-occurrence with PMOS. The presence of symptoms such as hair loss, dry skin, and bladder incontinence points to systemic involvement extending beyond the reproductive tract, potentially reflecting the interplay of hormonal dysregulation, immune dysfunction, and chronic inflammation characteristic of both conditions (1,43). These findings highlight the value of prospectively logged symptom data in capturing the full breadth of disease burden, and suggest that individuals with co-occurring endometriosis and PMOS may benefit from symptom-specific clinical attention beyond standard gynecologic care.

### Strengths and Limitations

With 1,872 participants reporting both endometriosis and PMOS, the AWHS offers a considerably larger and more population-representative sample than prior studies relying on smaller clinical samples of women with infertility or undergoing laparoscopy. Beyond broad survey-based data, prospective mobile symptom logging provided insights into menstrual symptom burden and minimized recall bias. Key limitations include potential surveillance bias, whereby diagnosed gynecologic conditions and infertility may reinforce each other’s detection, potentially leading to a relative overestimation of infertility burden across condition groups. More broadly, individuals with two diagnoses may have undergone more intensive clinical workups, potentially contributing to greater detection of unrelated conditions and the higher comorbidity burden observed in the endometriosis+PMOS group. All conditions were based on self-reported physician diagnoses, introducing potential misclassification. Cross-validation with CHR data demonstrated consistently higher self-reported than clinically confirmed prevalences, which may reflect both over-reporting and underdiagnosis in clinical records. As individuals typically accumulate multiple health records over their lifetime, the available CHR data likely capture an incomplete diagnostic history, contributing to lower concordance with self-reported conditions. Self-reported endometriosis and PMOS have been shown to be valid proxies for population-based research (44,45), and our prevalence estimates are consistent with population-based literature (4,46,47), lending support to their validity. As symptom logs reflect both physiological experience and app engagement (48), true symptom absence could not be distinguished from non-logging, potentially underestimating symptom burden. However, as this limitation applies equally across groups, its impact on between-group differences is likely minimal. Finally, overrepresentation of non-Hispanic White and highly educated individuals in the AWHS cohort (26) limits generalizability to the broader US menstruating population.

## Conclusions

In this large, population-based US cohort, co-occurring endometriosis and PMOS affected 2% of participants and was associated with a poorer health profile than either condition alone across reproductive and clinical domains. Individuals with endometriosis+PMOS had markedly elevated infertility odds reflecting compounded risk beyond the sum of individual condition effects, alongside the highest burden of comorbid mental and physical health conditions, the poorest subjective health, and a distinct menstrual symptom profile. Clinicians should be aware of this high-risk phenotype and consider evaluating for both conditions in individuals presenting with features of either. Dedicated research into the sequence of condition onset, specific risk factors, and long-term health trajectories is needed to inform clinical guidelines and improve care for this population.

## Data Availability

Aggregated data that support the findings of this study may be available upon reasonable request to the corresponding author. Any request for data will be evaluated and responded to in a manner consistent with policies intended to protect participant confidentiality and language in the study protocol and informed consent form.

## Acknowledgment

The AWHS team would like to thank the study participants for consenting and contributing to the advancement of women’s health research. We would also like to thank Carrie Sarcione and Kayley Abrams for their project management support.

## List of abbreviations

AWHS: Apple Women’s Health Study
BMI: Body mass index
CHR: Clinical health record
PMOS: Polyendocrine metabolic ovarian syndrome
RERI: Relative excess risk due to interaction
SES: Socioeconomic status
US: United States

## References

1. Teede HJ, Khomami MB, Morman R, Laven JSE, Joham AE, Costello MF, et al. Polyendocrine metabolic ovarian syndrome, the new name for polycystic ovary syndrome: a multistep global consensus process. The Lancet. 2026 May 12;0(0). doi:10.1016/S0140-6736(26)00717-8

2. Practice Committee of the American Society for Reproductive Medicine. Endometriosis and infertility: a committee opinion. Fertil Steril. 2012 Sep;98(3):591–8. doi:10.1016/j.fertnstert.2012.05.031 PubMed PMID: 22704630.

3. Schliep KC, Ghabayen L, Shaaban M, Hughes FR, Pollack AZ, Stanford JB, et al. Examining the co-occurrence of endometriosis and polycystic ovarian syndrome. AJOG Glob Rep. 2023 Aug;3(3):100259. doi:10.1016/j.xagr.2023.100259 PubMed PMID: 37663310; PubMed Central PMCID: PMC10472311.

4. Neven ACH, Forslund M, Ranasinha S, Sethi P, Dhungana RR, Mousa A, et al. Prevalence of polycystic ovary syndrome: a global and regional systematic review and meta-analysis. Hum Reprod Update. 2026 May 1;32(3):277–312. doi:10.1093/humupd/dmaf030 PubMed PMID: 41528735.

5. Teede HJ, Tay CT, Laven JJE, Dokras A, Moran LJ, Piltonen TT, et al. Recommendations from the 2023 international evidence-based guideline for the assessment and management of polycystic ovary syndrome. Eur J Endocrinol. 2023 Aug 2;189(2):G43–64. doi:10.1093/ejendo/lvad096 PubMed PMID: 37580861.

6. Stener-Victorin E, Teede H, Norman RJ, Legro R, Goodarzi MO, Dokras A, et al. Polycystic ovary syndrome. Nat Rev Dis Primer. 2024 Apr 18;10(1):27. doi:10.1038/s41572-024-00511-3 PubMed PMID: 38637590.

7. Cooney LG, Dokras A. Beyond fertility: polycystic ovary syndrome and long-term health. Fertil Steril. 2018 Oct;110(5):794–809. doi:10.1016/j.fertnstert.2018.08.021 PubMed PMID: 30316414.

8. Zondervan KT, Becker CM, Missmer SA. Endometriosis. N Engl J Med. 2020 Mar 26;382(13):1244–56. doi:10.1056/NEJMra1810764

9. De Corte P, Klinghardt M, von Stockum S, Heinemann K. Time to Diagnose Endometriosis: Current Status, Challenges and Regional Characteristics—A Systematic Literature Review. Bjog. 2025 Jan;132(2):118–30. doi:10.1111/1471-0528.17973 PubMed PMID: 39373298; PubMed Central PMCID: PMC11625652.

10. Madsen ME, Hartwell D, Kähler LKA, Dyrved L, Ejsing BH, Vexø LE, et al. Patients with endometriosis in the vagina, bowel, or bladder experience a prolonged diagnostic delay: an observational study. Hum Reprod. 2025 May 1;40(5):834–42. doi:10.1093/humrep/deaf046

11. Gibson-Helm M, Teede H, Dunaif A, Dokras A. Delayed Diagnosis and a Lack of Information Associated With Dissatisfaction in Women With Polycystic Ovary Syndrome. J Clin Endocrinol Metab. 2016 Dec 1;102(2):604–12. doi:10.1210/jc.2016-2963 PubMed PMID: 27906550; PubMed Central PMCID: PMC6283441.

12. Dinsdale NL, Crespi BJ. Endometriosis and polycystic ovary syndrome are diametric disorders. Evol Appl. 2021 May 14;14(7):1693–715. doi:10.1111/eva.13244 PubMed PMID: 34295358; PubMed Central PMCID: PMC8288001.

13. Dinsdale N, Nepomnaschy P, Crespi B. The evolutionary biology of endometriosis. Evol Med Public Health. 2021 Mar 12;9(1):174–91. doi:10.1093/emph/eoab008 PubMed PMID: 33854783; PubMed Central PMCID: PMC8030264.

14. Ismail S, Majdell C, Badgheish A, Baghlaf H, Dahan MH. Pregnancy, delivery, and neonatal outcomes among women with PCOS and endometriosis: a population database cohort. Arch Gynecol Obstet. 2024 Aug;310(2):1235–43. doi:10.1007/s00404-024-07589-w PubMed PMID: 38861027.

15. Sadeghi MR. Polycystic Ovarian Syndrome and Endometriosis as Two Evil Extremes of Health Continuum. J Reprod Infertil. 2022;23(1):1–2. doi:10.18502/jri.v23i1.8445 PubMed PMID: 36045878; PubMed Central PMCID: PMC9361725.

16. Crespi BJ, Evans SF. Prenatal Origins of Endometriosis Pathology and Pain: Reviewing the Evidence of a Role for Low Testosterone. J Pain Res. 2023 Feb 3;16:307–16. doi:10.2147/JPR.S389166 PubMed PMID: 36762368; PubMed Central PMCID: PMC9904225.

17. Ramezani Tehrani F, Noroozzadeh M, Zahediasl S, Piryaei A, Hashemi S, Azizi F. The Time of Prenatal Androgen Exposure Affects Development of Polycystic Ovary Syndrome-Like Phenotype in Adulthood in Female Rats. Int J Endocrinol Metab. 2014 Apr 1;12(2):e16502. doi:10.5812/ijem.16502 PubMed PMID: 24910644; PubMed Central PMCID: PMC4030220.

18. Dumesic DA, Abbott DH, Padmanabhan V. Polycystic Ovary Syndrome and its Developmental Origins. Rev Endocr Metab Disord. 2007 Jun;8(2):127–41. doi:10.1007/s11154-007-9046-0 PubMed PMID: 17659447; PubMed Central PMCID: PMC2935197.

19. Abbara A, Dhillo WS. Targeting Elevated GnRH Pulsatility to Treat Polycystic Ovary Syndrome. J Clin Endocrinol Metab. 2021 Jun 12;106(10):e4275–7. doi:10.1210/clinem/dgab422 PubMed PMID: 34117885; PubMed Central PMCID: PMC8475200.

20. Kumar P, Sait SF. Luteinizing hormone and its dilemma in ovulation induction. J Hum Reprod Sci. 2011;4(1):2–7. doi:10.4103/0974-1208.82351 PubMed PMID: 21772731; PubMed Central PMCID: PMC3136063.

21. McCartney CR, Marshall JC. Polycystic Ovary Syndrome. N Engl J Med. 2016 Jul 7;375(1):54–64. doi:10.1056/NEJMcp1514916

22. Stamatiades GA, Carroll RS, Kaiser UB. GnRH—A Key Regulator of FSH. Endocrinology. 2018 Dec 4;160(1):57–67. doi:10.1210/en.2018-00889 PubMed PMID: 30517625; PubMed Central PMCID: PMC6304106.

23. Mistry DS, Tsutsumi R, Fernandez M, Sharma S, Cardenas SA, Lawson MA, et al. Gonadotropin-releasing hormone pulse sensitivity of follicle-stimulating hormone-beta gene is mediated by differential expression of positive regulatory activator protein 1 factors and corepressors SKIL and TGIF1. Mol Endocrinol. 2011 Aug;25(8):1387–403. doi:10.1210/me.2011-0032 PubMed PMID: 21659477; PubMed Central PMCID: PMC3146254.

24. Bulun SE, Yilmaz BD, Sison C, Miyazaki K, Bernardi L, Liu S, et al. Endometriosis. Endocr Rev. 2019 Aug 1;40(4):1048–79. doi:10.1210/er.2018-00242

25. Holoch KJ, Savaris RF, Forstein DA, Miller PB, HigdonIII HL, Likes CE, et al. Coexistence of Polycystic Ovary Syndrome and Endometriosis in Women with Infertility. J Endometr Pelvic Pain Disord. 2014 Apr 1;6(2):79–83. doi:10.5301/je.5000181

26. Mahalingaiah S, Fruh V, Rodriguez E, Konanki SC, Onnela JP, de Figueiredo Veiga A, et al. Design and methods of the Apple Women’s Health Study: a digital longitudinal cohort study. Am J Obstet Gynecol. 2022 Apr;226(4):545.e1-545.e29. doi:10.1016/j.ajog.2021.09.041 PubMed PMID: 34610322; PubMed Central PMCID: PMC10518829.

27. Adnan T, Coull BA, Jukic AM, Mahalingaiah S. The real-world applications of the symptom tracking functionality available to menstrual health tracking apps. Curr Opin Endocrinol Diabetes Obes. 2021 Dec;28(6):574–86. doi:10.1097/MED.0000000000000682

28. Moss RH, Kelly B, Bird PK, Pickett KE. Examining individual social status using the MacArthur Scale of Subjective Social Status: Findings from the Born in Bradford study. SSM -Popul Health. 2023 Sep;23:101463. doi:10.1016/j.ssmph.2023.101463 PubMed PMID: 37691981; PubMed Central PMCID: PMC10492156.

29. Gibson EA, Li H, Fruh V, Gabra M, Asokan G, Jukic AMZ, et al. Covid-19 vaccination and menstrual cycle length in the Apple Women’s Health Study. Npj Digit Med. 2022 Nov 2;5(1):165. doi:10.1038/s41746-022-00711-9

30. Li H, Gibson EA, Jukic AMZ, Baird DD, Wilcox AJ, Curry CL, et al. Menstrual cycle length variation by demographic characteristics from the Apple Women’s Health Study. NPJ Digit Med. 2023 May 29;6(1):100. doi:10.1038/s41746-023-00848-1 PubMed PMID: 37248288; PubMed Central PMCID: PMC10226714.

31. Hager M, Wenzl R, Riesenhuber S, Marschalek J, Kuessel L, Mayrhofer D, et al. The Prevalence of Incidental Endometriosis in Women Undergoing Laparoscopic Ovarian Drilling for Clomiphene-Resistant Polycystic Ovary Syndrome: A Retrospective Cohort Study and Meta-Analysis. J Clin Med. 2019 Aug 14;8(8):1210. doi:10.3390/jcm8081210 PubMed PMID: 31416144; PubMed Central PMCID: PMC6722764.

32. Kim EJ, Kim T, Conigliaro J, Liebschutz JM, Paasche-Orlow MK, Hanchate AD. Racial and Ethnic Disparities in Diagnosis of Chronic Medical Conditions in the USA. J Gen Intern Med. 2018 Jul;33(7):1116–23. doi:10.1007/s11606-018-4471-1 PubMed PMID: 29736755; PubMed Central PMCID: PMC6025658.

33. Vela MB, Erondu AI, Smith NA, Peek ME, Woodruff JN, Chin MH. Eliminating Explicit and Implicit Biases in Health Care: Evidence and Research Needs. Annu Rev Public Health. 2022 Apr 5;43:477–501. doi:10.1146/annurev-publhealth-052620-103528 PubMed PMID: 35020445; PubMed Central PMCID: PMC9172268.

34. Rojas J, Chávez M, Olivar L, Rojas M, Morillo J, Mejías J, et al. Polycystic Ovary Syndrome, Insulin Resistance, and Obesity: Navigating the Pathophysiologic Labyrinth. Int J Reprod Med. 2014;2014:719050. doi:10.1155/2014/719050 PubMed PMID: 25763405; PubMed Central PMCID: PMC4334071.

35. Hong J, Yi KW. What is the link between endometriosis and adiposity? Obstet Gynecol Sci. 2022 May;65(3):227–33. doi:10.5468/ogs.21343 PubMed PMID: 35081675; PubMed Central PMCID: PMC9119730.

36. Lu MY, Niu JL, Liu B. The risk of endometriosis by early menarche is recently increased: a meta-analysis of literature published from 2000 to 2020. Arch Gynecol Obstet. 2023 Jan;307(1):59–69. doi:10.1007/s00404-022-06541-0 PubMed PMID: 35377041.

37. Nnoaham KE, Webster P, Kumbang J, Kennedy SH, Zondervan KT. Is early age at menarche a risk factor for endometriosis? A systematic review and meta-analysis of case-control studies. Fertil Steril. 2012 Sep;98(3):702-712.e6. doi:10.1016/j.fertnstert.2012.05.035 PubMed PMID: 22728052; PubMed Central PMCID: PMC3502866.

38. Tinano FR, Machado IFR, Latronico AC, Gomes LG. Shared Pathophysiological Mechanisms and Genetic Factors in Early Menarche and Polycystic Ovary Syndrome. J Neurosci. 2025 Mar 12;45(11):e1681242024. doi:10.1523/JNEUROSCI.1681-24.2024 PubMed PMID: 40074331; PubMed Central PMCID: PMC11905354.

39. Ma Y, Cai J, Liu LW, Hou W, Wei Z, Wang Y, et al. Age at menarche and polycystic ovary syndrome: A Mendelian randomization study. Int J Gynecol Obstet. 2023;162(3):1050–6. doi:10.1002/ijgo.14820

40. Lim SS, Norman RJ, Davies MJ, Moran LJ. The effect of obesity on polycystic ovary syndrome: a systematic review and meta-analysis. Obes Rev Off J Int Assoc Study Obes. 2013 Feb;14(2):95–109. doi:10.1111/j.1467-789X.2012.01053.x PubMed PMID: 23114091.

41. Allaire C, Bedaiwy MA, Yong PJ. Diagnosis and management of endometriosis. CMAJ Can Med Assoc J. 2023 Mar 14;195(10):E363–71. doi:10.1503/cmaj.220637 PubMed PMID: 36918177; PubMed Central PMCID: PMC10120420.

42. Dason ES, Koshkina O, Chan C, Sobel M. Diagnosis and management of polycystic ovarian syndrome. CMAJ Can Med Assoc J. 2024 Jan 29;196(3):E85–94. doi:10.1503/cmaj.231251 PubMed PMID: 38286488; PubMed Central PMCID: PMC10833093.

43. Petraglia F, Vannuccini S, Donati C, Jeljeli M, Bourdon M, Chapron C. Endometriosis and comorbidities: molecular mechanisms and clinical implications. Trends Mol Med. 2026 May 1;32(5):484–98. doi:10.1016/j.molmed.2025.09.002

44. Shafrir AL, Wise LA, Palmer JR, Shuaib ZO, Katuska LM, Vinayak P, et al. Validity of self-reported endometriosis: a comparison across four cohorts. Hum Reprod Oxf Engl. 2021 Apr 20;36(5):1268–78. doi:10.1093/humrep/deab012 PubMed PMID: 33595055; PubMed Central PMCID: PMC8366297.

45. Chan JL, Pall M, Ezeh U, Mathur R, Pisarska MD, Azziz R. Screening for Androgen Excess in Women: Accuracy of Self-Reported Excess Body Hair Growth and Menstrual Dysfunction. J Clin Endocrinol Metab. 2020 May 22;105(10):e3688–95. doi:10.1210/clinem/dgz264 PubMed PMID: 32442282; PubMed Central PMCID: PMC7448931.

46. Cox CM, Thoma ME, Tchangalova N, Mburu G, Bornstein MJ, Johnson CL, et al. Infertility prevalence and the methods of estimation from 1990 to 2021: a systematic review and meta-analysis. Hum Reprod Open. 2022 Jan 1;2022(4):hoac051. doi:10.1093/hropen/hoac051

47. Wang MH, Chen JH, Qi XY, Li ZX, Huang Y. Global prevalence of adenomyosis and endometriosis: a systematic review and meta-analysis. Reprod Biol Endocrinol. 2025 Nov 19;23(1):148. doi:10.1186/s12958-025-01483-z

48. Li K, Urteaga I, Wiggins CH, Druet A, Shea A, Vitzthum VJ, et al. Characterizing physiological and symptomatic variation in menstrual cycles using self-tracked mobile-health data. NPJ Digit Med. 2020;3:79. doi:10.1038/s41746-020-0269-8 PubMed PMID: 32509976; PubMed Central PMCID: PMC7250828.

