## Supplementary Material for "Menstrual symptoms and infertility odds among individuals with co-occurring endometriosis and polyendocrine metabolic ovarian syndrome"

### Supplement

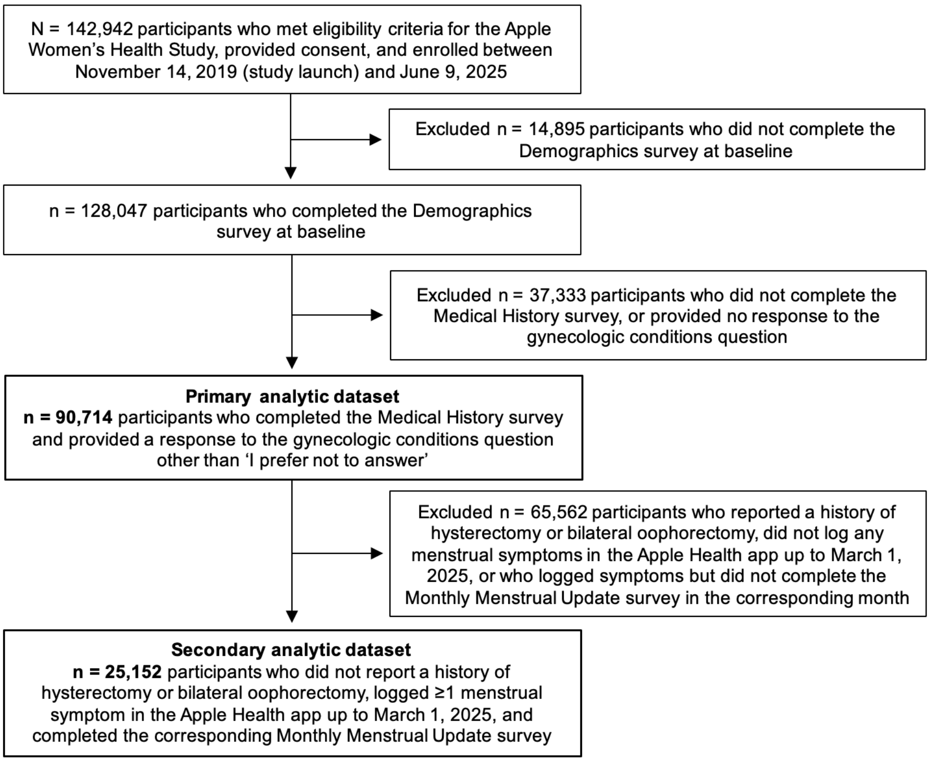


**Supplemental figure 1. Flow diagram of participants included in the analytic datasets.**

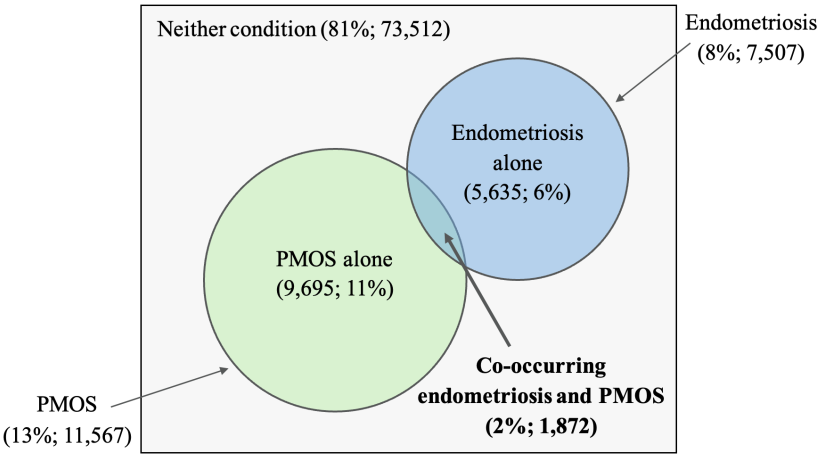


**Supplemental figure 2. Venn diagram illustrating the prevalence and overlap of endometriosis and polyendocrine metabolic ovarian syndrome (PMOS) in the analytic sample (N = 90,714).** Circle sizes are not drawn to scale.

**Supplemental table 1. Reproductive history, medical history, lifestyle characteristics, and reasons for menstrual cycle logging at baseline by condition group.**

| | Study sample (n = 90,714) | PMOS alone (n = 9,695) | Endometriosis alone (n = 5,635) | Endometriosis & PMOS (n = 1,872) | Neither condition (n = 73,512) |
|---|---|---|---|---|---|
| **Age at menarche in years,** n (%) | | | | | |
| ≤11 (early menarche) | 26,857 (29.6) | 3,369 (34.7) | 1,763 (31.3) | 725 (38.7) | 21,000 (28.6) |
| 12-15 | 55,905 (61.6) | 5,423 (55.9) | 3,386 (60.1) | 981 (52.4) | 46,115 (62.7) |
| ≥16 (late menarche) | 2,885 (3.2) | 434 (4.5) | 218 (3.9) | 88 (4.7) | 2,145 (2.9) |
| *Missing* | *5,067 (5.6)* | *469 (4.8)* | *268 (4.8)* | *78 (4.2)* | *4,252 (5.8)* |
| **Time to cycle regularity since menarche**, n (%) | | | | | |
| <1 year | 31,447 (34.7) | 2,224 (22.9) | 2,113 (37.5) | 462 (24.7) | 26,648 (36.2) |
| 1-2 years | 15,608 (17.2) | 1,088 (11.2) | 899 (16.0) | 240 (12.8) | 13,381 (18.2) |
| ≥3 years | 8,983 (9.9) | 1,049 (10.8) | 550 (9.8) | 214 (11.4) | 7,170 (9.8) |
| Regular after using hormones | 10,713 (11.8) | 2,085 (21.5) | 712 (12.6) | 314 (16.8) | 7,602 (10.3) |
| Not yet regular | 8,588 (9.5) | 2,168 (22.4) | 510 (9.1) | 433 (23.1) | 5,477 (7.5) |
| *Missing* | *15,375 (16.9)* | *1,081 (11.2)* | *851 (15.1)* | *209 (11.2)* | *13,234 (18.0)* |
| **Gravidity**, n (%) | | | | | |
| 0 | 37,341 (41.2) | 4,305 (44.4) | 1,511 (26.8) | 548 (29.3) | 30,977 (42.1) |
| 1 | 11,973 (13.2) | 1,480 (15.3) | 785 (13.9) | 290 (15.5) | 9,418 (12.8) |
| 2-3 | 22,665 (25.0) | 2,275 (23.5) | 1,809 (32.1) | 509 (27.2) | 18,072 (24.6) |
| ≥4 | 13,628 (15.0) | 1,162 (12.0) | 1,244 (22.1) | 429 (22.9) | 10,793 (14.7) |
| *Missing* | *5,107 (5.6)* | *473 (4.9)* | *286 (5.1)* | *96 (5.1)* | *4,252 (5.8)* |
| **Parity**, n (%)[a] | | | | | |
| 0 | 46,540 (51.3) | 5,409 (55.8) | 2,122 (37.7) | 823 (44.0) | 38,186 (51.9) |
| 1 | 12,619 (13.9) | 1,565 (16.1) | 935 (16.6) | 345 (18.4) | 9,774 (13.3) |
| 2–3 | 23,150 (25.5) | 2,062 (21.3) | 1,959 (34.8) | 541 (28.9) | 18,588 (25.3) |
| ≥4 | 4,442 (4.9) | 292 (3.0) | 389 (6.9) | 96 (5.1) | 3,665 (5.0) |
| *Missing* | *3,963 (4.4)* | *367 (3.8)* | *230 (4.1)* | *67 (3.6)* | *3,299 (4.5)* |
| **Hormone use for menstrual cycle management, ever, n (%)** | | | | | |
| Yes | 41,902 (46.2) | 6,604 (68.1) | 3,479 (61.7) | 1,377 (73.6) | 30,442 (41.4) |
| No | 44,575 (49.1) | 2,692 (27.8) | 1,917 (34.0) | 423 (22.6) | 39,543 (53.8) |
| *Missing* | *4,237 (4.7)* | *399 (4.1)* | *239 (4.2)* | *72 (3.8)* | *3,527 (4.8)* |
| **Hormone use to increase likelihood of pregnancy, ever**, n (%) | | | | | |
| Yes | 4,945 (5.5) | 1,520 (15.7) | 537 (9.5) | 365 (19.5) | 2,523 (3.4) |
| No | 81,532 (89.9) | 7,776 (80.2) | 4,859 (86.2) | 1,435 (76.7) | 67,462 (91.8) |
| *Missing* | *4,237 (4.7)* | *399 (4.1)* | *239 (4.2)* | *72 (3.8)* | *3,527 (4.8)* |
| **Family history of polyendocrine metabolic ovarian syndrome**, n (%) | | | | | |
| Yes | 6,985 (7.7) | 2,253 (23.2) | 429 (7.6) | 553 (29.5) | 3,750 (5.1) |
| No | 82,085 (90.5) | 7,277 (75.1) | 5,095 (90.4) | 1,279 (68.3) | 68,434 (93.1) |
| *Missing* | *1,644 (1.8)* | *165 (1.7)* | *111 (2.0)* | *40 (2.1)* | *1,328 (1.8)* |
| **Family history of endometriosis**, n (%) | | | | | |
| Yes | 8,603 (9.5) | 1,067 (11.0) | 1,692 (30.0) | 666 (35.6) | 5,178 (7.0) |
| No | 80,467 (88.7) | 8,463 (87.3) | 3,832 (68.0) | 1,166 (62.3) | 67,006 (91.1) |
| *Missing* | *1,644 (1.8)* | *165 (1.7)* | *111 (2.0)* | *40 (2.1)* | *1,328 (1.8)* |
| **Gynecologic conditions, ever diagnosed**, n (%) | | | | | |
| Endometriosis | 7,507 (8.3) | - | - | - | - |
| Polyendocrine metabolic ovarian syndrome | 11,567 (12.8) | - | - | - | - |
| Abnormal Pap smear | 24,645 (27.2) | 2,757 (28.4) | 2,208 (39.2) | 794 (42.4) | 18,886 (25.7) |
| Uterine fibroids | 7,724 (8.5) | 889 (9.2) | 1,343 (23.8) | 558 (29.8) | 4,934 (6.7) |
| Premenstrual syndrome/Premenstrual dysphoric disorder | 12,507 (13.8) | 1,970 (20.3) | 1,673 (29.7) | 735 (39.3) | 8,129 (11.1) |

| | | | | | |
|---|---|---|---|---|---|
| **Infertility, ever diagnosed, n (%)** | | | | | |
| Infertility overall | 4,676 (5.2) | 1,400 (14.4) | 728 (12.9) | 518 (27.7) | 2,030 (2.8) |
| *Primary infertility*[b] | *2,032 (43.5)* | *640 (45.7)* | *321 (44.1)* | *234 (45.2)* | *837 (41.2)* |
| *Secondary infertility*[c] | *2,484 (53.1)* | *713 (50.9)* | *379 (52.1)* | *269 (51.9)* | *1,123 (55.3)* |
| *Missing information on parity* | *160 (3.4)* | *47 (3.4)* | *28 (3.8)* | *15 (2.9)* | *70 (3.4)* |
| **Gynecologic surgeries**, n (%) | | | | | |
| Hysterectomy | 4,738 (5.2) | 374 (3.9) | 1,415 (25.1) | 430 (23.0) | 2,519 (3.4) |
| Unilateral oophorectomy | 1,902 (2.1) | 281 (2.9) | 486 (8.6) | 172 (9.2) | 963 (1.3) |
| Bilateral oophorectomy | 1,527 (1.7) | 114 (1.2) | 514 (9.1) | 190 (10.1) | 709 (1.0) |
| Removal or ligation of one fallopian tube | 2,085 (2.3) | 242 (2.5) | 372 (6.6) | 130 (6.9) | 1,341 (1.8) |
| Removal or ligation of both fallopian tubes | 8,905 (9.8) | 879 (9.1) | 1,277 (22.7) | 487 (26.0) | 6,262 (8.5) |
| **Endocrine conditions, ever diagnosed**, n (%) | | | | | |
| Hyperprolactinemia | 1,080 (1.2) | 275 (2.8) | 83 (1.5) | 79 (4.2) | 643 (0.9) |
| Hyperthyroidism | 2,876 (3.2) | 383 (4.0) | 300 (5.3) | 144 (7.7) | 2,049 (2.8) |
| Hypothyroidism | 9,797 (10.8) | 1,581 (16.3) | 915 (16.2) | 407 (21.7) | 6,894 (9.4) |
| Vitamin D deficiency | 27,169 (30.0) | 4,220 (43.5) | 2,290 (40.6) | 947 (50.6) | 19,712 (26.8) |
| **Cardiovascular conditions, ever diagnosed**, n (%) | | | | | |
| Coronary artery disease | 363 (0.4) | 45 (0.5) | 47 (0.8) | 33 (1.8) | 238 (0.3) |
| High cholesterol | 12,210 (13.5) | 1,820 (18.8) | 1,121 (19.9) | 445 (23.8) | 8,824 (12.0) |
| Hypertension | 11,524 (12.7) | 1,758 (18.1) | 1,080 (19.2) | 474 (25.3) | 8,212 (11.2) |
| Stroke | 629 (0.7) | 62 (0.6) | 63 (1.1) | 47 (2.5) | 457 (0.6) |
| Transient ischemic attack | 914 (1.0) | 112 (1.2) | 132 (2.3) | 81 (4.3) | 589 (0.8) |
| **Blood conditions, ever diagnosed**, n (%) | | | | | |
| Anemia | 26,875 (29.6) | 3,166 (32.7) | 2,509 (44.5) | 950 (50.7) | 20,250 (27.5) |
| Deep vein thrombosis | 1,409 (1.6) | 181 (1.9) | 165 (2.9) | 89 (4.8) | 974 (1.3) |
| Pulmonary embolism | 912 (1.0) | 141 (1.5) | 112 (2.0) | 60 (3.2) | 599 (0.8) |
| Thrombophilia | 388 (0.4) | 59 (0.6) | 40 (0.7) | 41 (2.2) | 248 (0.3) |
| **Digestive conditions, ever diagnosed**, n (%) | | | | | |
| Crohn's disease or ulcerative colitis | 902 (1.0) | 114 (1.2) | 110 (2.0) | 54 (2.9) | 624 (0.8) |
| Irritable bowel syndrome | 13,764 (15.2) | 2,008 (20.7) | 1,580 (28.0) | 688 (36.8) | 9,488 (12.9) |
| **Neurological conditions, ever diagnosed**, n (%) | | | | | |
| Chronic fatigue syndrome | 4,619 (5.1) | 724 (7.5) | 636 (11.3) | 361 (19.3) | 2,898 (3.9) |
| Headaches | 23,651 (26.1) | 3,178 (32.8) | 2,272 (40.3) | 954 (51.0) | 17,247 (23.5) |
| Insomnia | 15,654 (17.3) | 2,152 (22.2) | 1,650 (29.3) | 719 (38.4) | 11,133 (15.1) |
| Migraine | 28,399 (31.3) | 3,836 (39.6) | 2,704 (48.0) | 1,064 (56.8) | 20,795 (28.3) |
| **Mental health conditions, ever diagnosed**, n (%) | | | | | |
| Anxiety disorder | 47,350 (52.2) | 5,871 (60.6) | 3,420 (60.7) | 1,327 (70.9) | 36,732 (50.0) |
| Depression | 47,802 (52.7) | 5,986 (61.7) | 3,453 (61.3) | 1,312 (70.1) | 37,051 (50.4) |
| Panic disorder | 10,945 (12.1) | 1,447 (14.9) | 1,065 (18.9) | 518 (27.7) | 7,915 (10.8) |
| **Infectious diseases, ever diagnosed**, n (%) | | | | | |
| Pelvic inflammatory disease | 2,019 (2.2) | 295 (3.0) | 334 (5.9) | 215 (11.5) | 1,175 (1.6) |
| Urinary Tract/Bladder Infections | 45,405 (50.1) | 5,391 (55.6) | 3,505 (62.2) | 1,231 (65.8) | 35,278 (48.0) |
| **Other conditions, ever diagnosed**, n (%) | | | | | |
| Asthma | 22,389 (24.7) | 2,876 (29.7) | 1,866 (33.1) | 780 (41.7) | 16,867 (22.9) |
| Fibromyalgia | 4,895 (5.4) | 746 (7.7) | 759 (13.5) | 348 (18.6) | 3,042 (4.1) |
| Ovarian cancer | 261 (0.3) | 34 (0.4) | 47 (0.8) | 38 (2.0) | 142 (0.2) |
| **Overall physical activity level**, n (%) (multiple choice) | | | | | |
| None | 2,783 (3.1) | 338 (3.5) | 178 (3.2) | 90 (4.8) | 2,177 (3.0) |
| Light | 35,919 (39.6) | 4,525 (46.7) | 2,319 (41.2) | 862 (46.0) | 28,213 (38.4) |
| Moderate | 39,993 (44.1) | 4,201 (43.3) | 2,646 (47.0) | 812 (43.4) | 32,334 (44.0) |
| Vigorous | 23,481 (25.9) | 1,837 (18.9) | 1,260 (22.4) | 358 (19.1) | 20,026 (27.2) |
| Strenuous | 2,889 (3.2) | 189 (1.9) | 143 (2.5) | 38 (2.0) | 2,519 (3.4) |

| | | | | | |
|---|---|---|---|---|---|
| **Any special diet (e.g., vegetarian, no gluten, no dairy),** yes (n, %) | 31,338 (34.5) | 3,807 (39.3) | 2,270 (40.3) | 791 (42.3) | 24,470 (33.3) |
| **Frequency of feeling overly sleeping during the day in past month,** (n, %) | | | | | |
| Never | 4,149 (4.6) | 299 (3.1) | 201 (3.6) | 54 (2.9) | 3,595 (4.9) |
| Rarely | 13,649 (15.0) | 1,144 (11.8) | 739 (13.1) | 162 (8.7) | 11,604 (15.8) |
| Sometimes | 28,251 (31.1) | 2,839 (29.3) | 1,572 (27.9) | 434 (23.2) | 23,406 (31.8) |
| Often | 24,212 (26.7) | 2,822 (29.1) | 1,546 (27.4) | 562 (30.0) | 19,282 (26.2) |
| Almost always | 17,407 (19.2) | 2,282 (23.5) | 1,392 (24.7) | 594 (31.7) | 13,139 (17.9) |
| *Missing* | *3,046 (3.4)* | *309 (3.2)* | *185 (3.3)* | *66 (3.5)* | *2,486 (3.4)* |
| **Frequency current alcohol use among individuals who have ever consumed alcohol,** (n, %)[d] | | | | | |
| Never | 9,174 (11.2) | 1,034 (11.7) | 774 (15.1) | 305 (17.8) | 7,061 (10.7) |
| Monthly or less | 29,498 (36.2) | 3,788 (42.9) | 1,899 (37.1) | 747 (43.7) | 23,064 (35.0) |
| 2-4 times/month | 18,578 (22.8) | 1,965 (22.3) | 1,046 (20.4) | 297 (17.4) | 15,270 (23.2) |
| 2-3 times/week | 10,753 (13.2) | 890 (10.1) | 583 (11.4) | 147 (8.6) | 9,133 (13.9) |
| 4+ times/week | 7,567 (9.3) | 538 (6.1) | 521 (10.2) | 120 (7.0) | 6,388 (9.7) |
| *Missing* | *6,028 (7.4)* | *615 (7.0)* | *295 (5.8)* | *94 (5.5)* | *5,024 (7.6)* |
| **Smoking status,** (n, %) | | | | | |
| Never smoker | 57,311 (63.2) | 6,089 (62.8) | 3,013 (53.5) | 974 (52.0) | 47,235 (64.3) |
| Active/current smoker | 10,550 (11.6) | 1,108 (11.4) | 920 (16.3) | 343 (18.3) | 8,179 (11.1) |
| Ever smoker | 18,315 (20.2) | 2,015 (20.8) | 1,436 (25.5) | 459 (24.5) | 14,405 (19.6) |
| *Missing* | *4,538 (5.0)* | *483 (5.0)* | *266 (4.7)* | *96 (5.1)* | *3,693 (5.0)* |
| **Smoking frequency among active smokers,** (n, %) | | | | | |
| Some days | 2,699 (25.6) | 305 (27.5) | 186 (20.2) | 80 (23.3) | 2,128 (26.0) |
| Every day | 7,734 (73.3) | 791 (71.4) | 722 (78.5) | 259 (75.5) | 5,962 (72.9) |
| *Missing* | *117 (1.1)* | *12 (1.1)* | *12 (1.3)* | *4 (1.2)* | *89 (1.1)* |
| **Frequency current marijuana use,** (n, %)[d] | | | | | |
| Not at all | 34,860 (56.2) | 3,680 (55.3) | 2,151 (55.2) | 692 (52.2) | 28,337 (56.6) |
| Some days | 15,311 (24.7) | 1,657 (24.9) | 952 (24.4) | 322 (24.3) | 12,380 (24.7) |
| Every day | 11,273 (18.2) | 1,257 (18.9) | 758 (19.4) | 294 (22.2) | 8,964 (17.9) |
| *Missing* | *530 (0.9)* | *57 (0.9)* | *37 (0.9)* | *17 (1.3)* | *419 (0.8)* |
| **Overall subjective health status,** (n, %) | | | | | |
| Much better | 8,373 (9.2) | 412 (4.2) | 506 (9.0) | 93 (5.0) | 7,362 (10.0) |
| Slightly better | 17,286 (19.1) | 1,224 (12.6) | 973 (17.3) | 228 (12.2) | 14,861 (20.2) |
| About the same | 30,110 (33.2) | 3,001 (31.0) | 1,710 (30.3) | 490 (26.2) | 24,909 (33.9) |
| Slightly worse | 20,414 (22.5) | 3,136 (32.3) | 1,359 (24.1) | 589 (31.5) | 15,330 (20.9) |
| Much worse | 5,543 (6.1) | 1,009 (10.4) | 607 (10.8) | 313 (16.7) | 3,614 (4.9) |
| *Missing* | *8,988 (9.9)* | *913 (9.4)* | *480 (8.5)* | *159 (8.5)* | *7,436 (10.1)* |
| **Reasons for menstrual cycle tracking,** (n, %)[e] (multiple choice) | | | | | |
| Avoiding pregnancy | 3,569 (24.5) | 388 (21.6) | 156 (21.7) | 52 (18.7) | 2,973 (25.3) |
| Curiosity | 6,984 (48.0) | 747 (41.6) | 297 (41.4) | 112 (40.3) | 5,828 (49.6) |
| Getting pregnant | 2,270 (15.6) | 445 (24.8) | 128 (17.8) | 78 (28.1) | 1,619 (13.8) |
| Having menstrual products available | 4,735 (32.6) | 493 (27.5) | 222 (30.9) | 85 (30.6) | 3,935 (33.5) |
| Planning events (e.g., vacation) | 4,458 (30.7) | 449 (25.0) | 200 (27.9) | 72 (25.9) | 3,737 (31.8) |
| Sharing with a doctor | 4,537 (31.2) | 749 (41.8) | 271 (37.7) | 149 (53.6) | 3,368 (28.7) |
| Timing medications | 728 (5.0) | 133 (7.4) | 60 (8.4) | 26 (9.4) | 509 (4.3) |
| Tracking symptoms | 7,337 (50.5) | 978 (54.5) | 420 (58.5) | 164 (59.0) | 5,775 (49.2) |
| Understanding cycle patterns | 9,856 (67.8) | 1,284 (71.6) | 460 (64.1) | 192 (69.1) | 7,920 (67.4) |

[a]Parity defined as the number of reported pregnancies lasting ≥24 weeks, derived from self-reported pregnancy history data
[b]Primary infertility defined as self-reported physician-diagnosed infertility with parity = 0, percentages are calculated among individuals with self-reported physician-diagnosed infertility only
[c]Secondary infertility defined as self-reported physician-diagnosed infertility with parity ≥ 1, percentages are calculated among individuals with self-reported physician-diagnosed infertility only
[d]Percentages refer to the subgroups of individuals who have ever consumed alcohol/marijuana as the denominator, rather than the total study population
[e]Percentages refer to the subgroup of individuals who have completed the menstrual cycle tracking survey at baseline as the denominator, rather than the total study population

**Supplemental table 2. Univariable mixed-effects logistic regression analysis of menstrual symptom logging per cycle by gynecologic condition group.** Reference category: neither condition.

| **Logged symptom** | **Polyendocrine metabolic ovarian syndrome alone** OR (95% CI) | **Endometriosis alone** OR (95% CI) | **Co-occurring endometriosis and polyendocrine metabolic ovarian syndrome** OR (95% CI) |
|---|---|---|---|
| Abdominal cramps | 0.98 (0.91–1.05) | 1.02 (0.92–1.13) | 0.84 (0.70–1.01) |
| Acne | 0.98 (0.90–1.07) | 0.90 (0.79–1.03) | 0.87 (0.69–1.11) |
| Appetite changes | 1.07 (0.97–1.18) | 1.08 (0.94–1.25) | 1.02 (0.79–1.33) |
| Bladder incontinence | 1.20 (0.99–1.46) | 1.74 (1.34–2.26) | 1.93 (1.23–3.02) |
| Bloating | 0.99 (0.91–1.08) | 1.08 (0.95–1.22) | 1.01 (0.81–1.25) |
| Breast pain | 1.03 (0.92–1.14) | 0.95 (0.81–1.11) | 1.17 (0.90–1.53) |
| Chills | 1.07 (0.92–1.24) | 1.58 (1.29–1.94) | 2.20 (1.57–3.07) |
| Constipation | 1.08 (0.96–1.22) | 1.27 (1.07–1.51) | 1.24 (0.91–1.68) |
| Diarrhea | 1.29 (1.17–1.42) | 1.10 (0.95–1.28) | 1.15 (0.88–1.49) |
| Dry skin | 1.11 (0.96–1.28) | 1.29 (1.05–1.59) | 2.11 (1.52–2.94) |
| Fatigue | 1.18 (1.08–1.28) | 1.31 (1.16–1.48) | 1.43 (1.16–1.77) |
| Fever | 1.19 (0.54–2.60) | 1.91 (0.74–4.94) | 1.87 (0.33–10.73) |
| Hair loss | 1.30 (1.03–1.64) | 1.71 (1.24–2.34) | 1.98 (1.15–3.42) |
| Headache | 1.03 (0.93–1.13) | 1.02 (0.89–1.17) | 1.02 (0.79–1.30) |
| Hot flashes | 1.17 (1.01–1.36) | 1.06 (0.85–1.34) | 1.33 (0.91–1.95) |
| Lower back pain | 1.11 (1.01–1.22) | 1.47 (1.28–1.68) | 1.21 (0.95–1.55) |
| Memory lapses | 1.13 (0.93–1.38) | 1.63 (1.26–2.11) | 1.53 (0.96–2.45) |
| Mood changes | 0.98 (0.90–1.08) | 0.91 (0.80–1.05) | 0.85 (0.67–1.09) |
| Nausea | 1.29 (1.16–1.42) | 1.45 (1.25–1.67) | 1.89 (1.48–2.42) |
| Night sweats | 1.00 (0.85–1.17) | 1.01 (0.80–1.27) | 1.08 (0.71–1.62) |
| Pelvic pain | 1.38 (1.22–1.56) | 2.50 (2.12–2.95) | 3.07 (2.32–4.05) |
| Sleep changes | 1.20 (1.07–1.34) | 1.01 (0.85–1.20) | 1.35 (1.01–1.80) |
| Vaginal dryness | 1.20 (0.99–1.47) | 1.39 (1.05–1.85) | 1.16 (0.67–1.99) |
| *Number of subjects* | *2,775* | *1,103* | *375* |
| *Number of cycles* | *16,507* | *7,271* | *2,033* |
| OR = odds ratio, CI = confidence interval | | | |
